# The patient voice: valproate, topiramate and MHRA regulation

**DOI:** 10.1101/2024.09.06.24313040

**Authors:** Jane Hanna, Faye Waddams, Sarah Jones, Rachel Arkell, Heather Angus-Leppan

**Author notes:** Funding: None.

## Abstract

Valproate and topiramate use are increasingly restricted by government regulations, such as those of the MHRA. The views of people with epilepsy and their families affected by these have been little studied. This qualitative study examines 19 people with epilepsy and their views on the restrictions, and examines the thematic outcomes of these views – in terms of direct damage of avoiding valproate or topiramate (including SUDEP), missed opportunities and disruption of belief systems. Recommendations to align the regulations to human rights and true informed decision making are made.

## Introduction

Epilepsy, migraine and in bipolar disease are effectively treated by Valproate, and in a recent survey 11% of women with epilepsy self-reported taking it [1]. It is the most effective medication for genetic generalised epilepsies (GGE), but is no longer listed as the first-line treatment for women [2]. The Pregnancy Prevention Programme (PPP) mandates “user independent” contraception (intrauterine devices or subdermal progesterone only implants) for women taking valproate. The MHRA introduced this regulation to reduce teratogenicity and/or developmental disorders in potential offspring of women taking it [3], projected as a 11% risk of birth defects and a 30–40% risk of neuro-developmental disabilities [4]. The MHRA’s stated aim is to prevent all pregnancies on valproate. In England, the recorded number of pregnant women prescribed valproate fell from 68 women (April - September 2018), to 17 women (October 2021-March 2022) [4]. There are no published data on the outcomes of these pregnancies, where dose-adjustment and other mitigations may have had an effect.

To date, the MHRA neither collects nor funds data collection on the harms of the new regulations in terms of breakthrough seizures, injuries, or deaths. The Confidential Enquiry into Maternal Deaths records a near doubling in epilepsy related maternal deaths [5, 6]. Ineffective medication regimes and lack of information about the risk to pregnant women and their unborn of sudden death are key causes of these deaths. MBRRACE reported in 2020 that the risk in deaths must be viewed against the backdrop of the introduction of PPP. A SUDEP Action survey of 2,158 women found most were unaware of potential for SUDEP and fatality and only 25% accessed pre-conception counselling clinics [7]. This is also a central theme from 16 bereaved families reporting deaths during pregnancy to the Epilepsy Deaths Register since 2016 [8].

Patients and their families have repeatedly raised concerns about the blanket nature of MHRA restrictions since before their introduction; speaking for all [9–13], and on behalf of those lacking the ability to have an informed consensual relationship especially those with severe mental health problems and intellectual disability [10, 12, 13].

Unintended and unmitigated harms increase as the MHRA restrictions extend, as predicted by SUDEP Action and others since 2016 [14]. The extension of valproate PPP MHRA regulation to all people younger than 55 years in November 2023 [15], makes UK regulations more extensive and different from most of the world [10–13] Removing the previous gender difference leaves more people vulnerable to inferior seizure and other symptom control [16, 17]. Critical source data used for this decision is withheld from patients, charities and health professionals despite multiple requests to the MHRA, including two Freedom of Information requests from amalgamated epilepsy charities led by SUDEP Action [18]. The MHRA records that “errors have been identified in the study that may impact on the results” requiring further analysis [15]. Based partly on this report that is not peer-reviewed, verified or published, a major regulatory shift was implemented, out of line with most of the world [19] and despite academic criticism of the lack of robust data [16.17]. UK professionals and charities face the impossible task of supporting people with epilepsy in informed decision-making without access to all of the information [15,18]. In 2024, it is a matter of grave concern that freedom of information requests from 2022 and legal challenges from women with epilepsy and from bereaved families remain outstanding. SUDEP Action has led multiple requests for national review and national scrutiny and an Oxfordshire County Council Joint Health and Overview Committee has now also requested this in January 2024 and again in June 2024 [23]. Recently PPP was extended to women <55 years taking topiramate [24], and there are MHRA plans to extend restrictions to other antiseizure medications [25].

The MHRA mandated information booklet quantifies maternal teratogenicity and neurodevelopmental disabilities in putative offspring, but not the benefits of valproate, comparisons with other potential treatments, the risks of uncontrolled epilepsy and SUDEP, nor the risks of “user independent” contraception. Despite calls from SUDEP Action supported by MBBRACE, there is no signposting of SUDEP risk and potential SUDEP mitigation, even in the face of knowledge that effective medication is the first defence against SUDEP [26]. Patients have no right of appeal against two clinicians declining to prescribe a life-saving drug, and clinicians are vulnerable to sanctions if VPA is prescribed without PPP. The MHRA have refused to include links to the SUDEP and Seizure Safety tools [27,28], as recommended by MBRRACE and NHS RightCare despite repeated requests from SUDEP Action, epilepsy patient and clinical organisations.

The SUDEP and Seizure Safety Checklist is a holistic check which includes well recognised risk factors associated with epilepsy fatality and mitigating measures e.g. 69% of SUDEP could be avoided if generalized tonic-clonic seizures (GTCS) were controlled because they carry a 27 fold increased risk [29]. SUDEP Action and others predicted a rise in deaths in women in pregnancy and their unborn during the consultation on the introduction of the Prevent programme in 2016 and advocated a holistic approach which included patients [30]. Despite this, there is scant prospective data on the effects of the changed regulation is being gathered [31]. Survey data [32] and population studies [33] predict 30-40% of women will have breakthrough seizures on withdrawing valproate and changing to a different medication (lamotrigine or levetiracetam).

Despite the major impact of these changes, and although the new Valproate measures have been phased, the patient voice remains largely unheeded. There is little data on the views of informed patients and families wishing to have the option of valproate or topiramate while maintaining the same reproductive choices as others in the community. This study gathers the views of patients, carers and families regarding the restrictions on valproate, and now topiramate, prescription who have been denied these choices.

## Methods

The sources of accounts presented below were the SUDEP Action Charity “Share your experience” Register, and patients participating in a valproate PPP service review, approved by the Royal Free London NHS Foundation Trust Governance service. The initial views were expressed to the charity or the clinician (consultant neurologist or epilepsy CNS) in response to discussion of the PPP and MHRA regulation between January 2022-June 2024. Statements from consenting people with epilepsy or their carers who wished to commence or continue valproate or topiramate, or from relatives of those who died where valproate may have prevented their deaths, were collected, summarised in results, and analysed.

Comments by some (Jo Stan Louise Becky Tess) were summarised by the clinician (HAL) from contemporaneous notes of verbal comments. Others gave a written account and these are recorded below. Two patients (SJ & FW) are co-authors, others have been given pseudonyms. Age ranges are given, and exact years of events, and other potentially identifiable specifics are removed.

Qualitative analysis was used to summarise themes of their statements. A thematic analysis, informed by Braun and Clarke [33,34] was completed. Themes were generated in accordance with what patients considered significant in sharing their stories, as interpreted by the authors.

## Results

**Table I.**
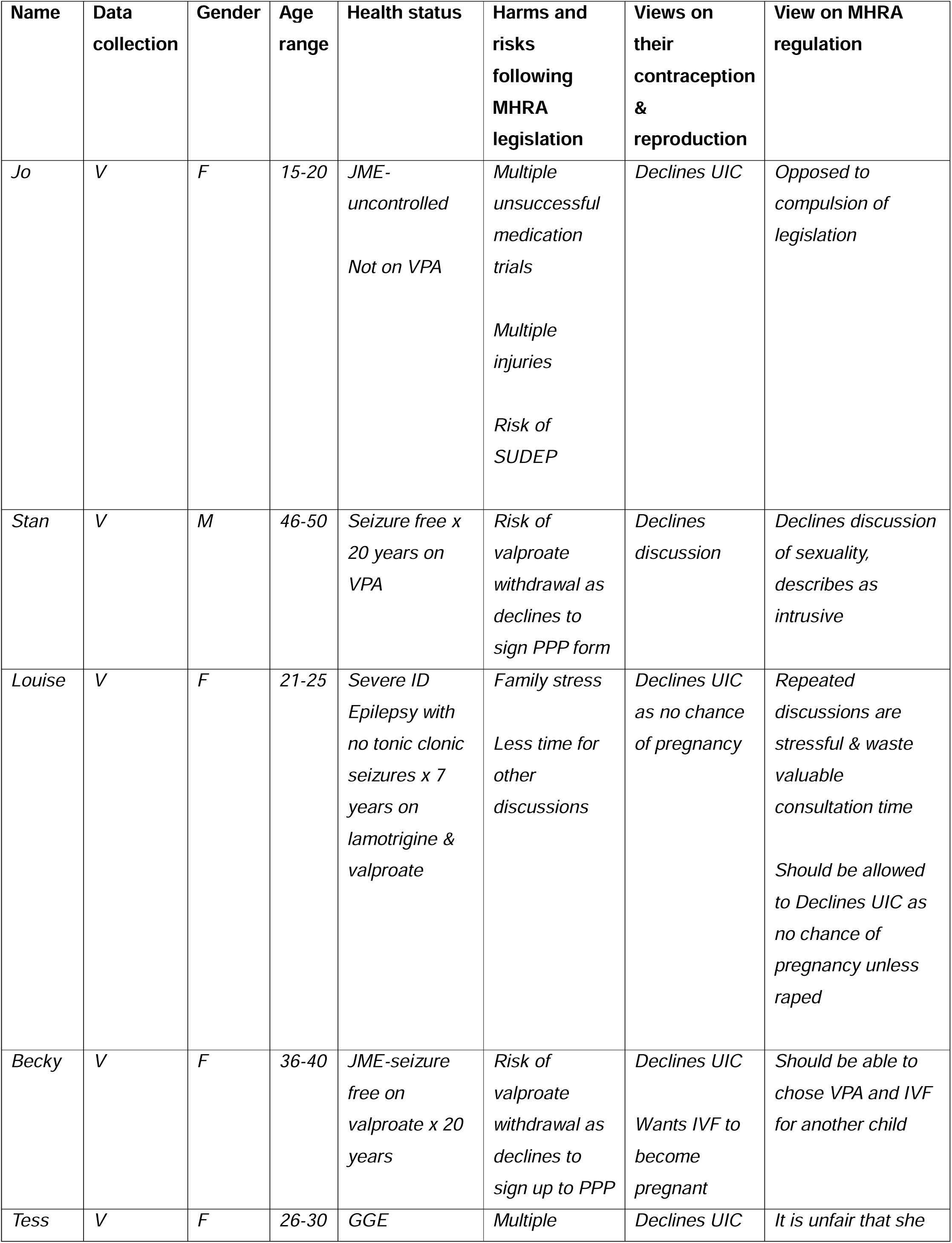
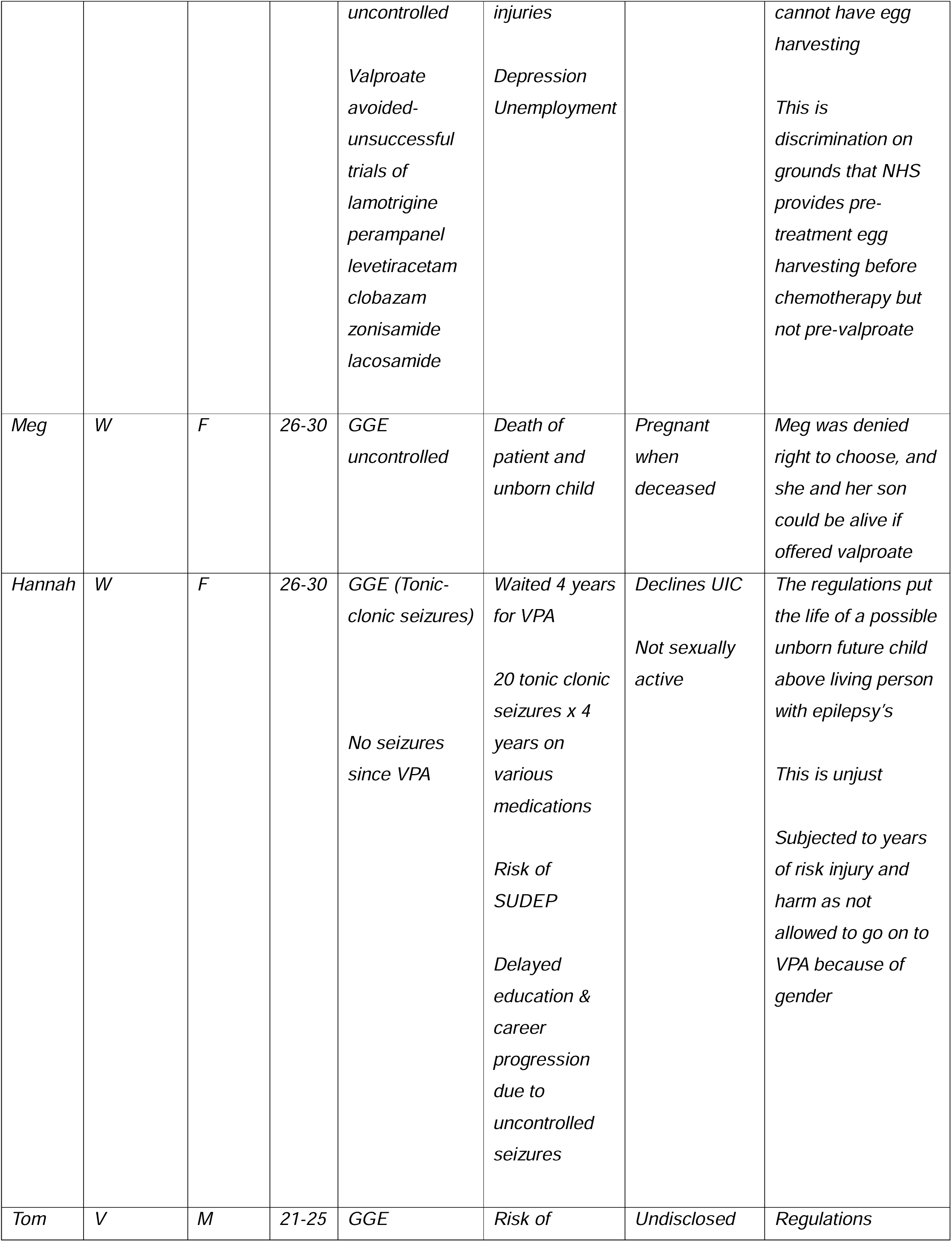
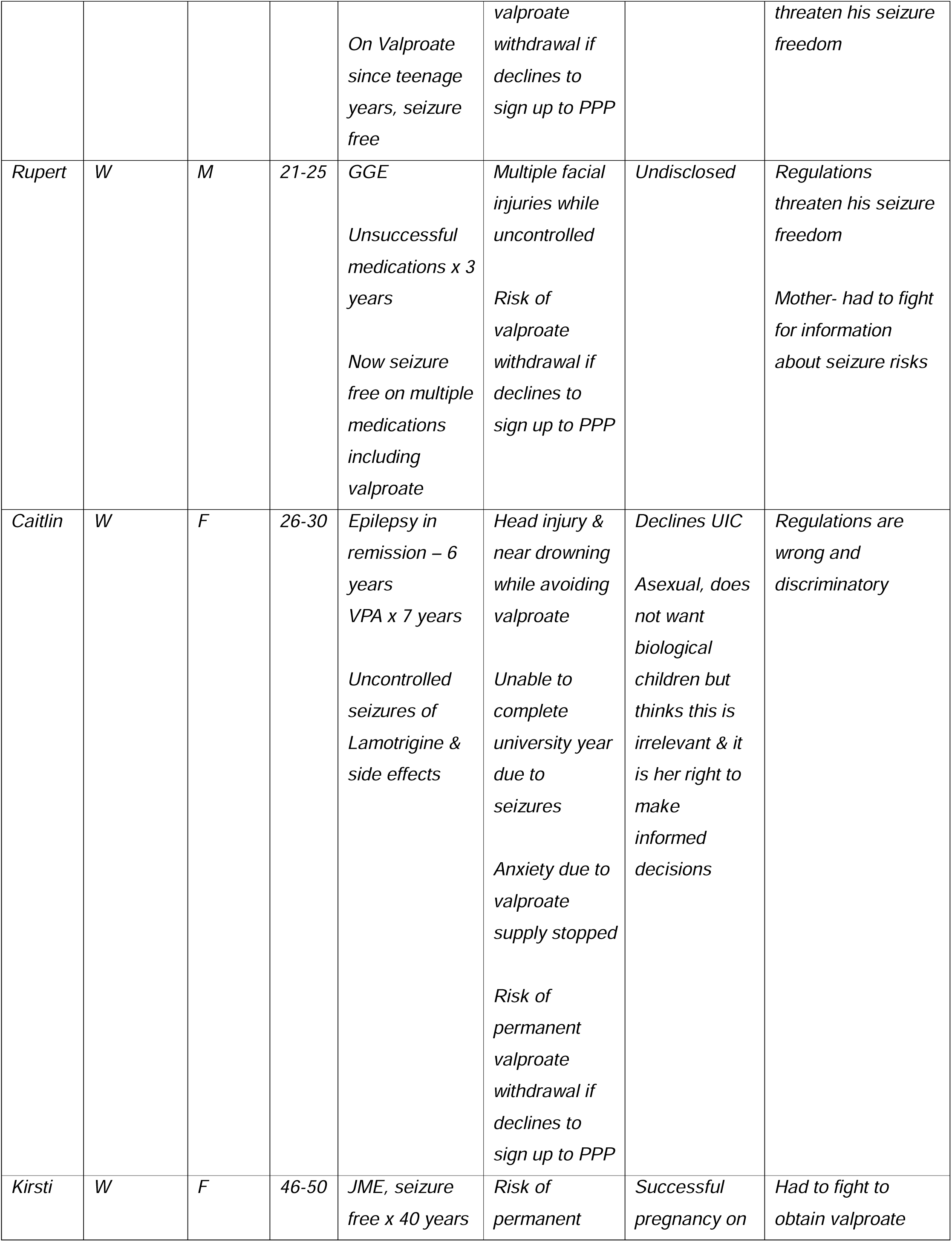
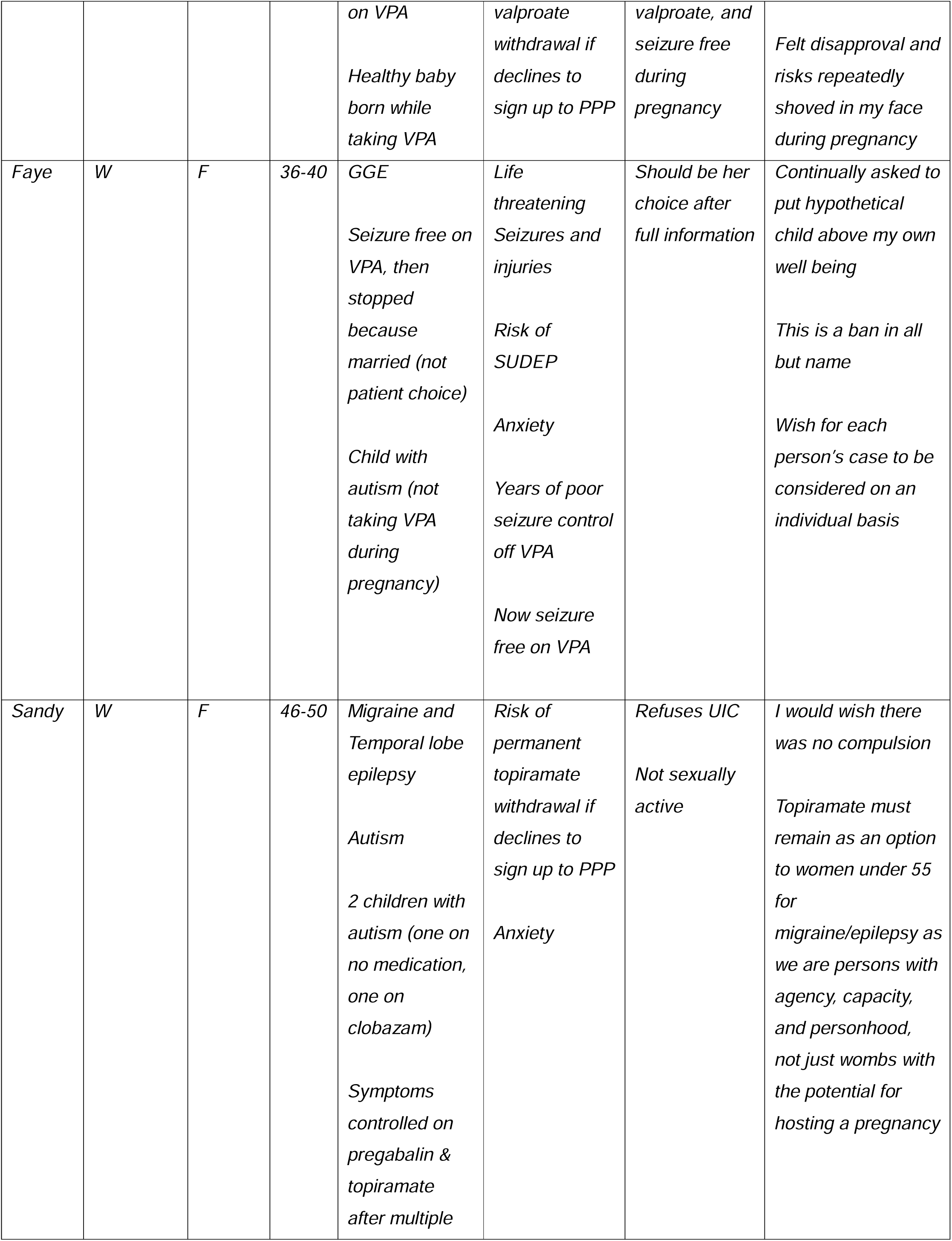
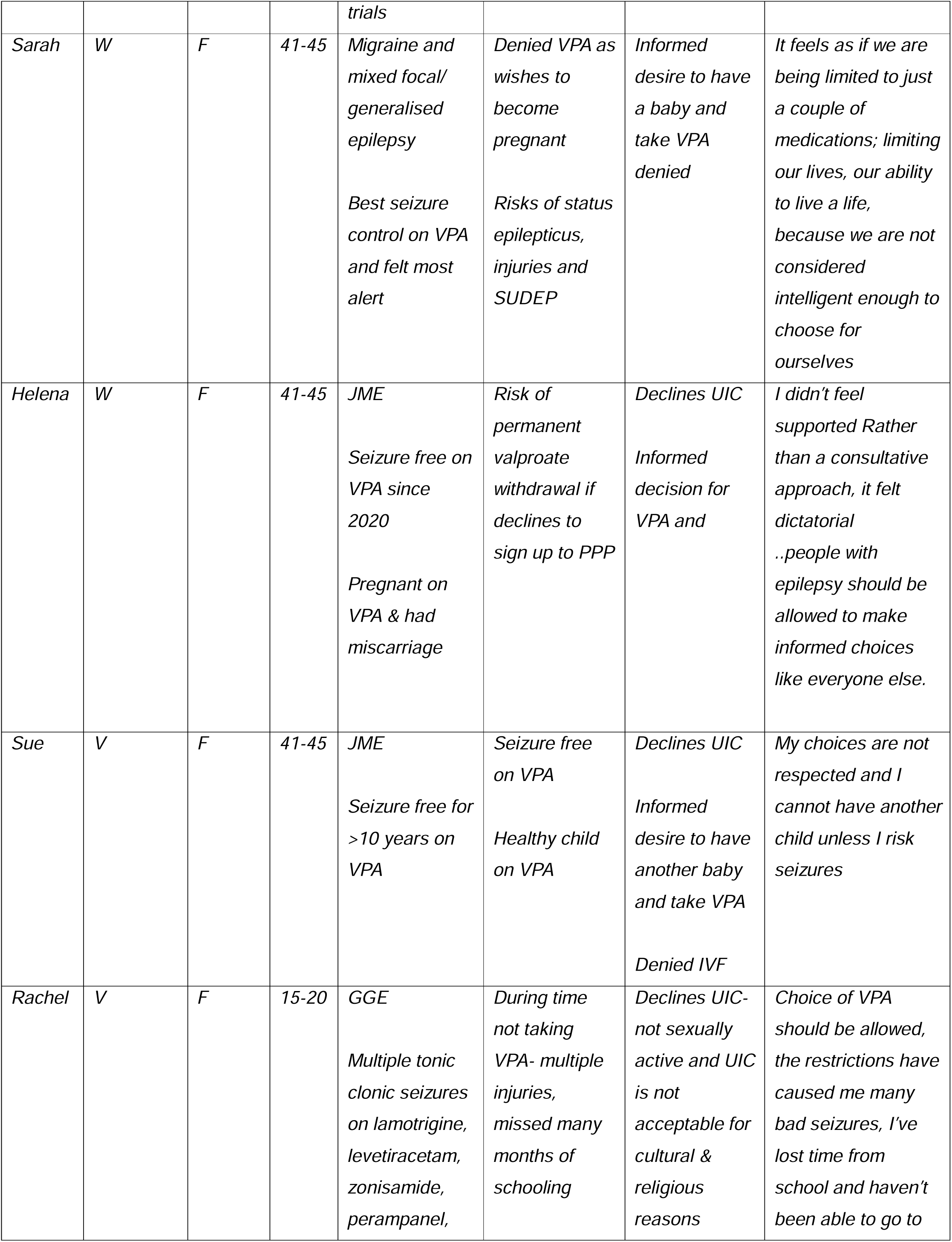
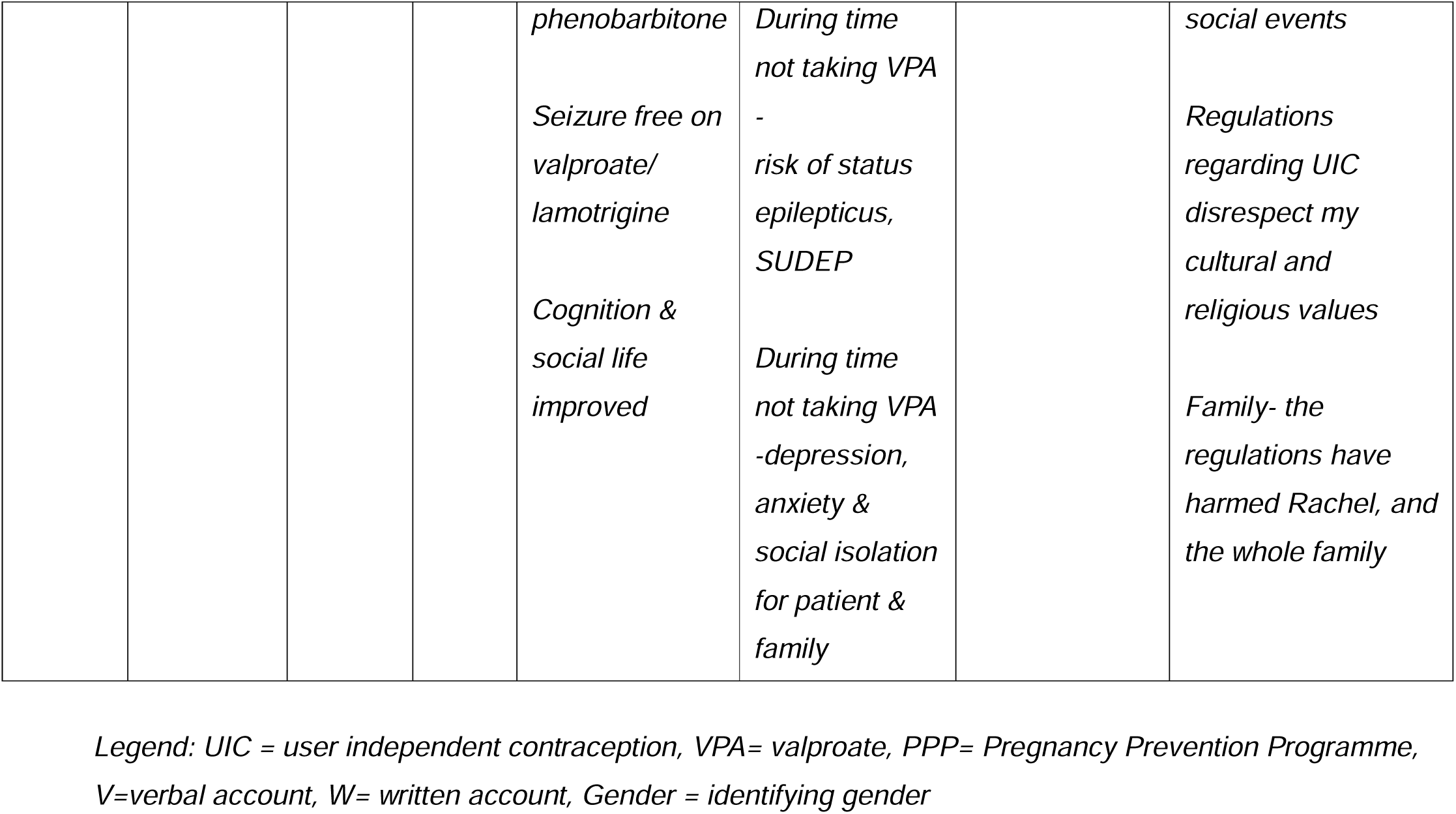
summarises the statements of patients and families.

### Meg and her family

A mother shares the tragedy of the sudden death of her daughter and her unborn grandchild. She is one of 19 suddenly bereaved families who are part of investigation by MBRRACE and are also participating in research with SUDEP Action on recent maternal deaths in epilepsy as part of the Epilepsy Deaths Research Register.

“My daughter was diagnosed with epilepsy when she was a young teenager. She was in her twenties when she and her unborn son died suddenly from a suspected epileptic attack. She was 17 weeks pregnant and had never been on sodium valproate.” Megan’s family think she could still be alive if her risk of SUDEP was properly assessed before and during her pregnancy, with discussions about her seizure management and medication. “I feel like she was not seen as an individual with epilepsy but in more general terms. Meg had the rest of her life ahead of her, we had an opportunity to be grandparents and all of that’s been stolen”

Alison is still waiting for an inquest seeking recognition of Meg’s Right to Life under Article 2 European Court Human Rights.

#### Hannah and her brother Tom

*“*The health of a future imaginary child seemed to be taken more seriously more than my life, they were receiving better care, they had more agency, and it was unjust”

*“*I have had twenty seizures over the last ten years, mostly generalised-tonic clonic seizures, that threaten my life. So many meds were tried at different doses for nearly four years. So many meds were tried at different doses for years. I was told that if I were a boy, I could start taking Sodium Valproate, which would be the most effective medication for me. There were concerns if, one day, I wanted to get pregnant - despite the fact that I’d never been interested in having children; I wasn’t sexually active; and I was only 16-20. The health of a future imaginary child seemed to be taken more seriously more than my life, they were receiving better care, they had more agency, and it was unjust.

I didn’t feel safe and I wasn’t safe either from SUDEP or from my mental health as by this time the community mental health team was visiting me every day. My depression started after my diagnosis, and my depression symptoms improving or worsening depending on whether my seizures improved.

I had to wait nearly four years to finally be offered Sodium Valproate, and I have not had a seizure on this medication. I was able to get on with life and am now healthy and have been training as a paramedic as part of my university degree. In contrast my brother was started on the drug straight away as a young teenager and was able to follow his dreams without his life being put on hold. Neither of us has any confidence that the new policy which accelerates PREVENT will give people with epilepsy the agency we need”

This month I was informed for the first time by my University some 18 months into the course that I could no longer fulfil my ambition to be a paramedic and experiencing the shock and impact of not having been told before.

#### Rupert

Like Hannah’s brother Tom Rupert needed Valproate to gain control of his seizures and has not been subject to the regime in the PREVENT programme for women, but could be this year if the next phase is introduced and would have been if he was newly diagnosed in 2024. For now there are the warnings about the risk to fertility which have been in medication packages since 2011. Caroline Scott shared her story as mother and a journalist https://sudep.org/article/mothers-perspective-journalist-caroline-scott

*“*My son Rupert, in his early 20s, had constant tonic clonic face-smashing seizures for over three years before a combination of drugs - different team at another hospital - helped get them under control last year. If I hadn’t understood his high risk of SUDEP I may not have pushed quite so hard. Days spent just trying to reach epilepsy nurse team, never mind the consultants. I know waiting lists a year long or more in other places”

#### Caitlin

“I have been recommended Valproate by two specialists and have been on it for seven years. I’ve been seizure-free for six years. Recently, I’ve had my valproate taken away unless I see another specialist who prescribes it yet again. I feel this is not only not in line with government recommendations for people continuing on Valproate but also incredibly disempowering for women.

My first seizure happened at night, and I spent the next day in A and E. For the next two years, I had uncontrolled seizures. Just before starting a university exchange overseas, I was prescribed Lamictal. I had further seizures. One resulted in a head injury and time in A and E. Another, I would have drowned had my mother not pulled me out of the water. The university placement was unsafe, and I returned home. As well as not controlling the seizures, Lamictal made me feel nauseous and depressed. In my early 20’s, I was prescribed Sodium Valproate.

I’m now in my late 20s and commuting, and the GP halted my drug with no warning. My mother had to take over communications with the GP as I work very long hours. It transpired that they stopped the Valproate with no warning because I don’t have a recent enough specialist, despite referring me to a specialist themselves weeks before. I have yet to go to that appointment, but me and my mother had to fight to get my Valproate back while I was running out. I’ve been promised a supply until I see the specialist, but after that, it’s up to him. I’m both asexual and do not want biological children, but I do not believe this should be relevant - I should be allowed to choose whether I take this drug and be trusted to take necessary precautions once informed of the risks.”

Caitlin has since been prescribed Valproate but she had to bring a verbal complaint.

#### Kristi and family

“I’ve had Photosensitive Juvenile Myoclonic Epilepsy / Generalised Idiopathic Epilepsy for 40 years now, I was diagnosed and I was prescribed Sodium Valproate. I count myself incredibly lucky, as this medication has been highly effective at preventing seizures for over 30 years now.

In the 2000s, my thoughts turned towards starting a family. I knew from the leaflet inside the box, I must talk to my doctor if thinking about getting pregnant, so over the years, I worked closely with my neurology consultant to understand all the risks and what measures I could do, to help minimize the risks down so I could make fully informed decision before starting a family. Due to pressing for all information concerning the risks and non-risks concerning Valproate, and after many lengthy conversations, I made the informed decision to remain upon the Valproate as the risks of coming off it and switching to another for myself and any future unborn, far outweighed the ‘potential’ risks to my future baby may or may not have.

When I fell pregnant, at the same time the forthcoming ‘Pregnancy Prevention Programme’ was being rolled out by the EMA and the MHRA concerning Valproate. Throughout my pregnancy, I had the ‘risks’ shoved into my face and a few times had medical professionals be disapproving towards myself when they saw what medication I was on. No woman should have to face this during an already stressful time, especially from medical ‘professionals’.

I gave birth to a very healthy, beautiful baby boy, who to this day continues to thrive and develop well. If I didn’t press for the balanced information nor ask how else the risks could be minimised so I could make an well balanced and informed decision with ALL FACTS, my son would not be here today, and/or myself if I’d taken the WRONG decision to come off from Sodium Valproate.

I strongly object to the ‘Pregnancy Prevention Programme’ and the awful language being used within the literature and graphics. I should not be compelled to have contraception and particular types of contraception to access a life-saving drug. I made an official complaint some years ago and have yet to have been given an answer to my complaint. If girls and women with epilepsy and now boys and men too are still to be allowed to have informed consent, the programme should be renamed as a RISK AWARENESS PROGRAMME with balanced resources (the national resources are not at all balanced); there need to be enough experts seeing people with epilepsy and GPs and pharmacists need to be supported to appreciate epilepsy risk. Otherwise many people with epilepsy will be deprived of their basic human rights and access to essential life saving medication and treatments because of state diktats and ignorance.

In 2024 Kristi is waiting for a second clinical signature so she can continue with Valproate. She has no right of appeal to anyone if this is refused and is experiencing ongoing anxiety about this. She first complained to the MHRA about the policy more than 7 years ago and her complaint is now with the Parliamentary Ombudsman.

#### Faye

“I was on valproate until I met my now husband in my 20s. My epilepsy nurse changed my medication. I have played the medication roulette ever since trying to find one that worked as well. I gave birth in the 2000s to my son. I had a difficult pregnancy and had seizures that risked both our lives. A decision was made then that he would be an only child. I carried on playing about with drug combinations to no success. I started to ask to go back on valproate. I got told no, I got told it was dangerous in pregnancy. Fine was my reply I’ll be sterilised to be met with questions about what If my husband wanted more children! My son is now at school and I have re trained as a lawyer. Children are still not in my plans and I wasn’t being listened to. I was continuously asked to put a hypothetical child above my own well being and that of my actual child. An idea that is absurd. In my late 30s and educated to masters level I am being told whats best for me. All I did whilst pregnant was try to protect my son and do what was best for him and now I was being denied the chance to do the same whilst he is here because of a child that will never be. Thats before you take account of my own wellbeing.

Eventually I was put back on Valproate after I was listened to. A number of weeks later I was in a car crash that left me trapped inside a vehicle for two hours on my head having to be cut out. Ten weeks on I would have assumed the stress, trauma and sleep deprivation along with pain would have caused seizures, however it’s been the biggest show that valproate is the right drug for me because I have not even had a hint of a seizure. My recovery has been slow and hard and I only dread to think what would have happened had we added seizures into the mix.

My wish is that this is not a blanket prevention. This is a ban in all but name, my wish is that each person’s case is considered on an individual basis and not based on the draconian measures in place that make it all but impossible to get the meds.”

#### Sandy

“I was in a car wreck in the 1990s, rear ended stationary at a red light. I was never the same. The migraines started almost immediately, and cost me my place at university, as I dropped out with severe pain.

And my first seizure was two years later. I will never forget sitting up from lying on the couch with terrible pain in my head as ever, startled with an inner rise, a disturbing feeling of apprehension of something wrong, and uttering a gibberish syllable. The next thing I knew, I was being hauled out on a stretcher, loaded into an ambulance, and being asked who the president was. I got the answer wrong by several years. A second seizure and several EEGs later, and a lifechanging diagnosis was given that saw me lose my driving licence and my freedom. My migraines were diagnosed formally and I was tried on loads of medications. My epilepsy was hit and miss after that, stabilised on lamotrigine after going through the gamut of phenytoin, valproic acid, and carbamazepine.

I moved to the UK in the 2000s. I did not have any more tonic clonics, but rather these strange “panic attacks” kept plaguing me. I ended up being told it was anxiety by my GP, given various benzodiazepines, counselling, propranolol for the migraines, and so on. My children were born, one unmedicated pregnancy and one whilst on clonazepam, and both were diagnosed with autistic spectrum disorder, as was my husband. My strange ‘panic attacks’ came and went, escalating at times of stress, and the migraines were a constant undercurrent of my life. Dark room, vomiting, photophobia, phonophobia, sleep required.

Life changed again for me when my strange ‘panic attacks’ that came and went entirely too quickly to be a panic attack (10 - 30 seconds) were diagnosed on the NHS at last as focal seizures (complex partial seizures was the term was at the time), and the migraines diagnosed formally again, after years of foboffery and time wasting at primary care level.

Lamotrigine coincided with a rash, so it was decided to switch meds. Topiramate was chosen because of its tag-team indications of epilepsy and migraine prevention. And combined with pregabalin, it helps me with both. I’ve been on it since. That’s about 12 years.

I was diagnosed with autism spectrum disorder myself about 10 years ago, not long after seeing the neurologist about epilepsy/migraine. My autistic children are autistic because of their parents, not because of the medications I have or haven’t taken.

I am now in my late 40s. I have been divorced. My children and I live on our own, disabled people forging a future in a country that seems hellbent on punishing us at every turn. My eldest has ASD, selective mutism, migraines, and hEDS. My youngest has ASD, ADHD, Tourettes, and ARFID.

I am utterly abstinent due to my circumstances. I’ve not been on birth control since before my eldest was born. I don’t want birth control, I didn’t even like HRT, it felt synthetic and wrong and harsh. Who would contraception be for? I have disabled children that occupy entirely all of me. While they are at school/university, I work In our spare time, i work hard to make sure my children are socialised, get out to do the things they like to do, get haircuts and make it to the opticians and the doctors and the older one can access her university things, the younger one can get to her drama performances and practice, and we go to anime cons and go to board gaming cafes and mix with people because otherwise, they would never go outside.

So, apologies for the novel, but finding this out about the topiramate PPP last friday took my breath away. I went through the gamut of feelings - anger that they’d be taking my choice away, abject fury that they think i’m a ‘risk’ to be managed just because i still have the plumbing, livid that a hypothetical foetus means more to the MHRA than my quality of life, my functioning, my ability to cope with the paperwork and admin of managing everything for my children and myself, all alone. I get no PIP for myself, it was refused, so I am literally left to struggle on alone, and I need my brain to function as best It can, to run this show. I feel the American right wing quasi-pregnancy stuff creeping into the UK and i worry deeply because I thought the UK was better than that. I left it all behind, you know?

Further reading all weekend led me to read the public document about the topiramate safety review and the fact that the MHRA appear to be de facto engaging in soft eugenics. Depo provera is not without harms, same with the coils of both kinds. Why do they think they have the right to force this choice onto women?

Here I am, a woman who decided no birth control, Why am i being forced to contemplate it now, just because I happen to still have a uterus?

They are in short trying to prevent autistic people being born. This is deeply problematic and if the autism charities knew this, it would be all over for the MHRA, it would absolutely explode.

How many of the women in the Bjork dataset are also autistic, like their children? Can the MHRA answer that? Kneejerk “shut it down” reactions from the regulator are not helpful - there are unintended consequences to denying women a useful medication just because they happen to have a womb and it’s a teratogen. Poor seizure control with a substitute medication, morbidity from migraine exacerbation, lost days at work, etc.

I’ve been hounded for months by the “pharmacy team” within my GP surgery for reasons I cannot get them to explain to me, I have no idea if it’s to do with topiramate, but if i lose it, i will be back to dark rooms, vomiting, sleeping it off, unreliability at my tiny little job, and being unable to fulfill my role for my children. I’ll be back to many more focal seizures, and that terrifies me. They’re not entirely controlled on the topiramate+pregabalin combo but I don’t want to go back to such severity that they generalise in my sleep again. Or i wake up in an ambulance again.

I would wish that there be no compulsion in any way. This ‘must’ only ever be a woman’s choice in line with long established medical ethics, not some misguided lawsuit / compensation prevention by the MHRA. Topiramate must remain available as an option to women under 55 for migraine/ epilepsy, as we are persons with agency, capacity, and personhood, not just wombs with the potential of hosting a pregnancy.”

#### Sarah

“I have been on so many medications now since being diagnosed with epilepsy a few years ago, with varying levels of impact. They are still constantly changing. Following a hospital stay last year due to being in Status, I was out on valproate for a few months - it was the lowest seizure rate I have ever had. It was wonderful, I hadn’t realised how much of my life I had missed due to seizures and also the side effects of the other meds (I had relatively little side effects from valproate).

But then I was taken off it: because I want to get pregnant. Now I’ve read the research, I’ve seen the rate of birth defects. I have spent my life risk assessing - whether for a company as CEO, in sports as a high-performance coach, when working with high level offenders and now, risk assessing the impact of IT systems in a healthcare setting. In all these situations, not only do I trust my ability to weigh up risks and make a decision based upon them, but other people (many thousands now) trust my ability and I do the same. Yet I am not allowed to risk assess my own choice to be on a medication and have a child. A child which after 7 miscarriages is, frankly, most likely a dream that will never happen.

Instead I am left on medications that do not work anywhere near as well. This when I think about it, I try not to for my sanity, leaves me at risk of injury from the amount of seizures I have or going into status again. As I and currently having 4-5 TC seizures a week, with no idea about the lesser seizures, this situation is probably a larger risk to my life than I want to think about. Plus the stress on my family and friends, the lack of confidence to go out and do activities that really matter to me because of the risk of having a seizure in public: all of these are having a very real, and large impact on my life. I found earlier this year that I had stopped doing anything that meant I might have a seizure in public, so wasn’t getting out of the house at all.

I do not want to take contraception; for my mental health I want to give it every chance try and have a child, I have to live for ever with the decision to basically not have a child; something the NHS does not provide any counselling (at all, let alone on a long term basis) for people with epilepsy making such a momentous decision. Plus the contraception itself comes with risks (possibly even greater), and if I’m really honest, being forced to be on contraception feels like eugenics.

I often feel that I can choose not to take my medication at all, but I can’t choose to take one that helps me. Why am being put in this situation? I have a condition that is made worse by stress, and yet the stress is being imposed by the lack of seizure control, and the decision to not take contraception. But above all, why I am not considered to be able to make a decision for myself?

We are told we can still get valproate by the charities and other government bodies: but in reality it’s not happening. Clinicians, rightly, seem to be scared to put patients on it. Instead we are subjected to constant medication adjustments trying to make another medication work better. And now to compound the situation, this fear seems to be spreading to other medications - including new ones, which were also relatively effective.

It’s strange - I can choose to drink or take drugs and have a child; I could be on medication for any other medical condition (including cancer meds) regardless of the fact that I am a woman. Yet with epilepsy these options are not open to me. It is discrimination at best, at worst people lives are being put at danger and no thought is being given to this it seems. There is certainly no support from NHS psychology services to assist with those of us left struggling, without a life, trying to decide if having a child really matters that much, being made to feel selfish for even daring to want a child.

It feels as if we are being limited to just a couple of medications; limiting our lives, our ability to live a life, because we are not considered intelligent enough to choose for ourselves. Epilepsy already makes you feel like a lesser person and shrinks your world beyond anything you could ever have imagined. This situation is making it even worse. What is it about epilepsy that is different to any other condition in terms of medication we can take, and choices we can make? We need to be treated as people, not as a condition, to at least be listened to, as we have not been given a voice. It seems no one is trying to hear us other than a very precious few.”

#### Helena

“I’m in my early 40s. I have worked in Recruitment/Talent Acquisition for global companies and perfectly capable of making my own decisions. I live on my own and have a driving licence.

I was diagnosed with Juvenile Myoclonic Epilepsy more than 25 years ago. I have had one tonic clonic seizure and also a few myoclonic jerks which occurred after a meningitis vaccination. I was prescribed Valproate and haven’t had seizures since then.

I have been very belt and braces over the years to prevent pregnancy previously being on the pill and using a barrier method. I also take 5 x the normal amount of folic acid, a high dose of vitamin D and multivitamins anyway. I thought condoms alone would be fine.

Over the decades of taking Valproate I have never been offered epilepsy pre- conception counselling. Whilst I had ideally over the years wanted a baby one day, I had resigned myself to the fact it wasn’t going to happen in light of changes to guidance on pregnancy in those taking sodium valproate. I had asked for detailed information over the years as to the risks, percentage of children born with disabilities vs without disabilities, the nature of what is classified as a disability and how the information was collected to decide if I would have children one day. This information was vague to the point it then became being advised the only way was to switch medication in advance of becoming pregnant.

I’ve also never been offered a SUDEP and Seizure Safety checklist either and only found out about EpsMon self-monitor App recently from someone else with epilepsy on social media. I’ve never had an opportunity for a balanced conversation about the benefits and risks of Valproate or the range of options I might have for treatment or if I wished to plan a family.

In March this year I unexpectedly became pregnant. On finding out I was pregnant I contacted my GP to request a scan to ascertain health and age of the foetus and get my head round how to proceed.

A scan was scheduled a few days later and my GP contacted the neurology team at the Hospital. My neurology nurse phoned me before the scan and was very clear that if I progressed with the pregnancy I’d be taken off valproate immediately and switched to Keppra, the health of the foetus could already be impacted and the risk of learning difficulties at 30-40% potentially higher.

The seriousness of my situation was stressed and I was advised to terminate. I did ask what if it was one of the ones that didn’t end up with a disability but the message was clear that the risks were high all round and keeping a child was only an option if I immediately changed medication. My neurology nurse appears reluctant for valproate to be used at all in pregnancy. It was very much a militant approach and like the end of the world.

I felt I had no real options but forced into following the outcome they wanted. At that point whilst time was important I had been looking to know the outcome of the scan and impartial information and advice to think through what myself and my partner wanted to do.

I then got the results of the scan from the pregnancy scanning unit which showed the foetus had stopped developing at a certain point and I would experience a miscarriage anyway which took place a few days later.

My neurology nurse panicked at the paperwork from the hospital which said I could try again for a child after my next period. I assume it was standard wording for that department and had to reassure my neurology nurse that I was not and would not be trying. He was happy with this. The following weeks were as stressful. I was told I would not be re prescribed valproate unless I went on a reliable contraceptive.

Their definition of reliable was the implant, Mirena coil or copper coil. Aside from that hysterectomy or sterilisation. I previously tried the Mirena coil and it didnt agree with me at all. I had clots on occasion the size of my hand despite not even being in a sexual relationship during that time.

I had to fight and persuade to get valproate prescribed again by stating I’d go on the pill even though I’m no longer in a relationship and would use condoms as well when I am. Fortunately I had some extra valproate and hadn’t run out when they changed my prescription to Keppra, otherwise I might have had a breakthrough seizure. It was this that made me so angry that they were prepared to switch my medication overnight and switched it whilst I was pregnant.

I didn’t feel supported and it felt like the agenda of everyone else was put before how I felt and what would impact my life. My feelings were clearly inconsequential. It was a very hard time. Rather than a consultative approach, it felt dictatorial and as long as I do what I’m told all is fine. It is very clear that if God forbid I have another contraceptive failure I will no longer receive valproate ever again.

I recognise that with recent campaigning by those who have children affected by valproate and didn’t receive information like I did, the NHS is wanting awareness and to not incur costs caused by lawsuits and compensation pay outs. However, I do feel it has gone the other way and people with epilepsy should be allowed to make informed choices like everyone else.

My neurology nurse appears reluctant for valproate to be used at all in pregnancy and Ive been told there is no researched safe dose. I’m so glad to hear these issues with PPP and the stringent approach is being discussed.”

### Thematic analysis

#### Key themes

The statements were analysed to generate key themes:

- Direct and indirect damages of avoiding Valproate and topiramate,
- the missed opportunities,
- disruption to belief systems of patients, and to their relationship with clinicians and the medical system as a whole.

#### Direct damage of avoiding Valproate and topiramate

The increased risk of seizures, injury, and in one tragic case death of mother and child, are highlighted in 9 out of the 17 cases. Increase in mental health problems (stress, anxiety and/or depression) for the person and families are highlighted in all cases.

#### Missed opportunities

Loss of life of the person and her unborn baby are tragically highlighted in Meg’s case. Loss of employment is specifically highlighted by Tess and Hannah, and academic loss is highlighted by Hannah, Caitlin, and Rachel. Loss of the full range of treatment choices is mentioned by all.

#### Disruption of belief systems

A stated lack of respect for their autonomy is highlighted in all people’s case histories.

This is highlighted by many of the voices as an existential feeling of loss of control of one’s life (Tess, Meg, Hannah, Tom, Rupert, Caitlin, Kirsty, Fay, Sandy, Sarah, Helena, Sue and Rachel). A sense of being disrespected is expressed, for example by Sarah, “It feels as if we are being limited to just a couple of medications: limiting our lives, our ability to live a life because we are not considered intelligent enough to choose for ourselves”.

Two people specifically mention that the regulations feel like an imposition of eugenics (Sandy and Sarah). The view of women as essentially and predominantly a vehicle for producing children, and men as a producer of healthy sperm is seen to underlie the MHRA regulation. The voices speak against this imposed belief system in all cases.

Many of the themes have echoes in the previous research, investigations and in advocacy for reform of regulations of Valproate and anti-seizure medication [36–39]. The true extent of damage in terms of increased seizure risk, injuries and death is uncertain as there is no systematic data, but surveys and the EURAP pregnancy register estimate that the risk of breakthrough seizures in women coming off Valproate is 30-40% [31,32].

Increased stress, anxiety and depression, and other mental health problems due to inability to obtain the medication the person with epilepsy wishes, has not been quantified in any scale. The scale of the effect of this on seizure control, and the risk of injuries, is unknown. Interpretations are also complicated by the fact that Valproate is a mood stabiliser, so changing to another medication may reduce mental health for that specific reason alone [16,17].

The theme of missed opportunities has been highlighted by researchers and advocates [36–39].

The imbalance of power and autonomy which the voices highlight has been reflected in previous writing [36–39]. The contrast between the choices available to a person with a genetic disorder in a society which purports to support diversity and informed decision-making is stark and has been highlighted [9, 11, 36–39]. For example, if a person has a genetic disease, with a 50% or greater chance of passing on this condition to a child born, the person has a right to have a child without any testing, have prenatal testing and a termination, or avoid having a child, or have IVF with an unaffected egg or sperm donor. Analogous choices are denied to a person who wishes to take Valproate. More subtle themes emerging are concern at the perception of an assumption underlying the regulation that if a woman or man is taking Valproate and has a child with disability, that that child’s life is not as valuable or worthy as other lives. The corollary is that if Valproate is avoided there is an assumption that the child will not have disabilities, which is not the case.

The people’s statements touch on many ethical concerns arise from these voices – firstly, the idea implied by the regulations that a disabled child has a life that is not of equal value to a child without disability. The need to find a source of blame for disability is also seen - if a disabled child is born from a parent taking Valproate, that that blame is external; whereas if the child is born with a genetic aetiology to a disability that is somehow the parent’s fault [36–38].

## Discussion

The views of patients and families who have suffered harms from restrictions on the availability of the antiseizure medications valproate and topiramate are little heard by policy makers.

This study goes some way to redressing this. It cannot reverse the deaths of those who could be alive if provided with effective anti-seizure medications. Their families want urgent learning from these deaths since 2016 and call for changes to be made including balanced policy making and communications and a precautionary approach to SUDEP and epilepsy deaths.

Against a context where SUDEP Action has worked with researchers and policy makers to bring to light a significant burden of sudden mortality in the young over two decades and warned in 2016 of rising maternal deaths and other harms, this neglect of patients and families is reckless to patient safety, patient rights, human dignity (36-39). The lived experience in this study appears to be the opposite of an experience informed by principles of better patient safety set out recently by the Patient Safety Commissioner: they did not experience a safety culture; being at the heart of considerations about their treatment; being treated as equals; nor were their risks clearly identified and mitigated. The one sided nature of the data collection on number of prescriptions for Valproate without regard to gathering data on harms to patients is unacceptable and extraordinary in a society championing equality of care.

Arising from the harms of the current valproate and topiramate regulations they support the campaign by SUDEP Action, clinicians, human rights lawyers and advocates and urgently call for:

- Suspension of January 2024 MHRA restrictions on valproate and topiramate pending further review national review and scrutiny of the PPP policy
- Reversion to ILAE 2016 guidelines
- Balanced written, online and verbal recording in guidelines of the impact of SUDEP and other harms including those of user independent contraception to allow informed individual and true consent
- Acknowledgement of the full range of patient lifestyle choices and sexual preferences in guidelines
- Revised MHRA and NHS information packs to include SUDEP and Seizure Safety Checklist and communication safety tools recommended by EMBRRACE and NHS RightCare
- Funding for additional clinical time for informed patient decision-making
- National publication of an impact assessment of the valproate and topiramate PPP on patient care and choice and on the local NHS of the PPP before the PPP comes into force
- Full disclosure of the CHM & MHRA proceedings regarding valproate and Full disclosure of the CHM & MHRA funding & Disclosures (individual & organisational) in the past, and in the future
- Government funded prospective registries of all maternal & foetal outcomes of all pregnancies in epilepsy and bipolar disease
- Government funded registry of patients switching from valproate to other medications
- Free and non-judgemental provision of all reproductive options (including egg harvesting, termination, surrogacy, adoption, sperm banking, abstinence, having a baby while taking valproate and topiramate) for people wishing to commence valproate or topiramate, and full acknowledgement of the diversity of human sexual choices (including straight, LGQBT+, ace)
- Protection of clinicians supporting informed choice of their patients from discrimination and prosecution

## Data Availability

All data produced in the present study are available upon reasonable request to the authors

## Acknowledgements

We thank the patients and families who have shared their stories to help others.

